# Impact of the Management Development Programme (MDP) on primary health care manager competencies and organisational Performance

**DOI:** 10.64898/2026.05.28.26354357

**Authors:** Tembeka Sineke, Khumbo Shumba, Aneesa Moolla, Constance Mongwenyana-Makhutle, Danleen Hongoro, Jacqui Miot, Petra Kruger, Jessie Graven, Dorina Onoya

**Affiliations:** Health Economics and Epidemiology Research Office, Department of Internal Medicine, School of Clinical Medicine, Faculty of Health Sciences, University of the Witwatersrand, Johannesburg, South Africa; Department of Work and Social Psychology, Maastricht University, Maastricht, the Netherlands; Aurum Institute, Cape Town, South Africa

**Author notes:** Correspondence: Dorina Onoya, PhD, 39 Empire Road, Health Economics and Epidemiology Research Office, Department of Internal Medicine, School of Clinical Medicine, Faculty of Health Sciences, University of the Witwatersrand, Johannesburg, South Africa.

## Abstract

Primary healthcare (PHC) managers are central to the functioning of South Africa’s healthcare system, yet many assume leadership roles without formal management training. To address this gap, the Aurum Institute developed the Management Development Programme (MDP), a structured leadership and management training intervention aimed at strengthening PHC management competencies. This study evaluated the impact of the MDP on leadership practices, organisational readiness for change, and workplace stress among PHC managers in the Western Cape Province. A non-randomised matched cluster trial was conducted across 20 PHC facilities. Intervention facilities were purposively selected based on participation in the MDP, while matched control facilities were randomly selected. Data were collected using structured and semi-structured surveys administered to facility managers and clinic staff. Leadership competency was assessed using the Leadership Practices Inventory (LPI), which measures five dimensions of exemplary leadership: Model the Way, Inspire a Shared Vision, Challenge the Process, Enable Others to Act, and Encourage the Heart. Organisational readiness for change was measured using Kotter’s 8-Step Framework, while workplace stress was assessed using a 13-item version of the Brief Job Stress Questionnaire focusing on Job Meaning, Environmental Quality, Autonomy, and Control. Intervention effects were estimated using generalised linear models adjusted for manager age, years in role, matched-pair fixed effects, and cluster-robust standard errors. Outcomes were reported as adjusted risk differences with 95% confidence intervals and two-sided p-values. A total of 20 facility managers (median age 51 years; IQR 42–55; 90% female) and 105 clinic staff members (median age 42 years; IQR 35–50) participated in the study. Managers in both intervention and control facilities reported consistently high self-rated leadership competency scores across all LPI domains, with no statistically significant differences between groups. Similarly, clinic staff rated managers highly across the standard LPI domains, and no significant differences were observed between intervention and control facilities. Despite the absence of significant differences in overall leadership competency scores, staff in intervention facilities reported significantly stronger relational and communication practices among managers compared with staff in control facilities (72.7% vs. 64.0%; adjusted risk difference 22.0%, 95% CI 6.1–37.8; p=.007). After adjustment for age and tenure imbalances, intervention facilities also demonstrated significantly higher scores for institutionalised capability and learning culture (adjusted risk difference 21.3%, 95% CI 0.6–42.0; p=.043). Managers who participated in the MDP further reported stronger perceptions of district support, including improved internal leadership and cultural readiness (adjusted risk difference 22.1%, 95% CI 14.0–30.3; p<.001) and greater district leadership and resource availability (adjusted risk difference 28.1%, 95% CI 15.6–40.6; p<.001). No statistically significant differences were observed in workplace stress across any domain. Although the MDP did not produce measurable short-term improvements in managers’ self-rated leadership competencies or standard LPI domains as assessed by staff, it was associated with important gains in relational leadership practices, organisational readiness for change, and perceived district support. These findings suggest that structured management training programmes may strengthen critical organisational and interpersonal foundations necessary for sustained performance improvement within PHC settings.

## INTRODUCTION

Primary healthcare clinics (PHCs) are integral to South Africa’s healthcare system, serving as the cornerstone of essential service delivery to communities. The 2019 General Household Survey (GHS) found that approximately 72.5% of households in South Africa rely on the public healthcare system as their primary source of healthcare services [1]. PHCs are the primary entry point for medical care and play a pivotal role in referring patients to specialized and hospital services [2]. However, South Africa’s healthcare system often faces many challenges, including resource constraints [3], which are further compounded by a lack of competent managers [3, 4]. Furthermore, health care systems are complex to manage and lead [5, 6]. Alongside other ongoing enhancements to primary healthcare quality, such as the Ideal Clinic Realisation and Maintenance (ICRM) program [7], recent evaluations have shown the importance of greater involvement and decision-making autonomy for PHC managers [7, 8]. Management of PHCs in South Africa has traditionally been assigned to senior, trained healthcare providers, such as professional nurses, who typically receive minimal training in facility management to address many challenges, including the scarcity of human resources [9, 10].

Recognising the need for a locally relevant training and mentoring opportunity for PHC managers, the Aurum Institute developed the Management Development Programme (MDP), initially targeting PHC managers but also supporting the development of managers at tertiary-level health facilities and other health units within the district departments of health (DOH)in South Africa. Since 2014, the MDP has been implemented across several provinces in South Africa, including Gauteng, Limpopo, North West, and Western Cape. While the programme continues to be rolled out in other provinces, there remains limited evidence on its effectiveness in improving PHC management practices. As national and provincial departments consider scaling up the MDP, evidence is needed to demonstrate its impact and inform decisions about broader implementation.

Evaluating leadership and management development programmes requires reliable and valid tools that are able to measure changes in leadership competencies and management practices. These tools are structured frameworks or instruments used to assess, guide, and improve organisational performance, leadership, decision-making, and team effectiveness[11]. The science behind management tools is rooted in organisational psychology, behavioural science, leadership theory, and psychometrics[12]. Effective management tools are typically designed to measure observable behaviours, competencies, or organizational processes reliably and validly, enabling organisations to identify strengths, gaps, and areas for development. In the current study, we apply a widely used leadership assessment tool, the Leadership Practices Inventory (LPI)[13, 14], to assess the competency skills of the clinic managers, and this was selected because it focuses on measurable leadership behaviours, making it particularly suitable for assessing competency development among PHC managers following structured leadership training.

This study aims to address this knowledge gap by evaluating the effectiveness of the MDP in strengthening PHC leadership. Specifically, we assess whether participation in the MDP improves the management competencies of PHC managers, compared to those who have not undergone the training. By examining the added value of structured leadership development, the study aims to generate evidence that can guide future programme investments and inform policy decision makers. This is particularly timely and important as South Africa moves towards implementing the proposed National Health Insurance (NHI) framework [15], where strong PHC leadership will be essential to ensure equitable and efficient healthcare delivery.

## MATERIALS AND METHODS

### Ethics statement

The study protocol was reviewed and approved by the Human Research Ethics Committee of the University of Witwatersrand (ETHICS REFERENCE NO: 240713). Written informed consent was obtained from all participants prior to their inclusion in the study. All personal identifiers were removed from the final analytic dataset.

### The Aurum Management Development Programme (MDP)

Since 2014, the Aurum Institute has partnered with Presidents’ Emergency Plan For AIDS Relief (PEPFAR), the South African National Department of Health (NDOH), and various provincial health departments to design, pilot and implement a customised management training to equip public sector PHC Managers with the skills to manage the day-to-day responsibilities involved in delivering safe and effective patient care, including management of human resources, finances, equipment and consumables, information and medical records. The intensive 12-month programme comprises:

- Classroom-based training on management theory and practices, conducted monthly for 2 days per month during the first year. Each module focuses on a different set of core competencies, such as operational planning, human resource management, infrastructure and facility organisation, and pharmacy management.
- One-on-one/small-group peer mentoring programme (during and after the formal training), comprising monthly workplace visits, off-site group mentoring, and/or virtual/telephonic support for participating managers to build their confidence in applying new skills.
- A portfolio of evidence comprising completed homework assignments that cover essential managerial tasks such as developing operational plans, conducting staff meetings, preparing facility reports, and implementing quality improvement projects. Not only does this ensure compliance with national standards, but it also provides the evidence to assess their competency for an accredited management qualification.

### Structure and conceptual framework for the evaluation

The evaluation employed both quantitative and qualitative methods. For the quantitative component, we determined the MDP’s impact on managers’ leadership skills, and their effects on PHC staff and patient care, including quality of care, patient satisfaction and specific health outcome metrics.

We applied Kotter’s eight-step change model, which is widely used to guide and evaluate organisational change management processes [16–18], including assessing the effectiveness of change initiatives, such as the Aurum MDP, in improving PHC managers’ skills, PHC staff experiences and patient experiences of quality of PHC care and outcomes. We hypothesised that a PHC manager who participated in the MDP would be better equipped to lead their staff through the steps outlined in Kotter’s model for organisational change.

Kotter’s model explains how an effective manager can overcome staff resistance to change and create a deeply ingrained, stable new way of working through eight steps. To improve clinic operational effectiveness by applying the skills developed in the MDP, the PHC manager has to prepare the clinic staff (organisation in this case) for change by breaking down the existing status quo, creating a sense of urgency for adopting new approaches (Step 1), motivating PHC staff to change by highlighting current problems and involving influential leaders in the change process (Step 2), and reducing barriers to change. The PHC manager has to create and communicate a clear vision (Steps 3 & 4), provide support and resources, and encourage participation and feedback (Steps 5 & 6). Consolidating Gains and Producing More Change: Anchoring New Approaches in the Culture (Steps 7 and 8).

### Study design overview by objectives

**Figure 1** illustrates the non-randomised, matched-cluster (PHC) trial design, in which PHCs whose managers were conveniently assigned to participate in the MDP program by the District authorities were compared with randomly selected matched controls, PHCs with no historical MDP support, to evaluate the impact of the MDP on PHC managers. The comparison facilities were matched by facility type, district, and, to a large extent, the subdistrict/area’s population density. The population density not only determines the burden of disease but also influences the size of the clinic’s patient population. In total participants were PHC managers and clinic staff from 20 PHCs in three districts in the Western Cape province of South Africa, including Garden Route (4 PHCs), Cape Winelands (6 PHCs), and the Cape Town Metropolitan (10 PHCs).

**Figure 1.**
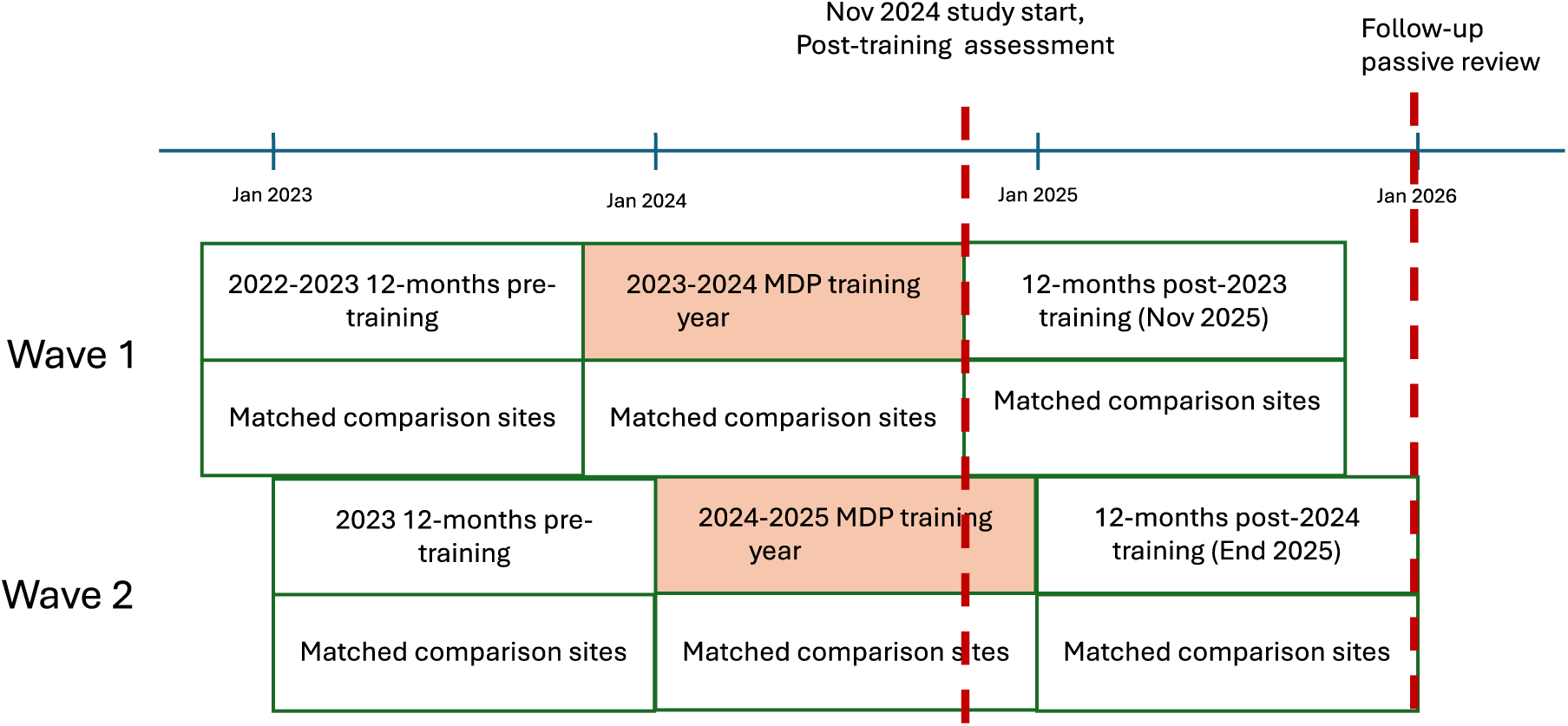
**Study Design**

### Study sites

For this study, the primary sampling unit was the PHC facility, its manager, with PHC staff and patients as secondary sampling units. MDP-supported clinics included PHC facilities whose managers participated in the MDP in 2023–2024 in the Western Cape Province (City of Cape Town Metro: n=5; Cape Winelands: n=2; Garden Route districts: n=3). MDP-supported clinics were selected and referred to the MDP by district leadership based on programmatic criteria (i.e., selection was determined by district management rather than self-selection by individual facility managers). Each MDP-supported clinic was matched to a comparison clinic that had no historical MDP support and was similar with respect to facility size and type and catchment population.

The study was organised into two waves to maximise evaluation of the MDP training cohorts enrolled in November 2023 (wave 1) and January 2024 (wave 2).

### Sample size

The study planned to include at least one PHC manager and five clinic staff per facility. From the MDP training cohorts, Wave 1, we targeted 5 managers, and Wave 2, we targeted 5 managers, giving a total of 10 managers in the intervention arm. An equal number of managers and staff were targeted in the control arm, giving a total sample size of 20 managers and 105 staff across both arms. We assumed that this sample size would be sufficient to gather different views on leadership.

### Participant Eligibility Criteria

Participants in this study included clinic managers (supporting managers where applicable) and clinic staff from selected primary healthcare (PHC) facilities in the intervention and comparison arms. Clinic managers or supporting managers were eligible if they were deployed at one of the study sites and held an active management role, such as facility manager or operational manager. For intervention sites, eligible managers must have participated in the MDP training in 2023 or later. All managers had to provide informed consent to participate and were determined not to have previously enrolled in the study at another site.

Clinic staff were eligible if they were actively working at the participating PHC during the study period and were willing to provide informed consent to participate in evaluating the leadership practices of their manager. Similarly, we did not include staff who previously enrolled in the study at another site. Only individuals with direct knowledge of their clinic manager’s leadership (typically from having worked under their supervision for at least three months) were invited to participate in the assessment.

### Data collection

The survey was conducted following the completion of the MDP training period (January–June 2025) for both training waves, at a point when the managers had completed all training modules and were actively applying components of the programme during the training year. Data collection involved PHC Managers from the intervention and control sites who were referred to the study with permission from the district health authorities. All consenting managers completed an interviewer-administered semi-structured survey to collect demographic information, work history, and experience in managing their PHC. Independent interviews were conducted with clinic staff to provide additional perspectives on the leadership competencies of their managers. Clinic staff were identified and enrolled, provided they were available at the time of the interview and met the inclusion criteria.

### Baseline pre-intervention interviews with PHC Managers and Staff

Managers and PHC staff from intervention clinics who completed their training year at the time of the study commencement, and pre-training information was collected at the post-intervention time point, before the period of independent skill application. Managers (n=20) from the intervention and comparison sites were approached independently after the Western Cape provincial research ethics board approved the study. Consenting managers completed an interviewer-administered semi-structured survey between January and March 2025 (at the end of 12-month MDP support for the intervention clinics) to collect demographic information, work experience, and experience in managing the PHC where they are currently posted. Interviews cover elements of Kotter’s 8-step change management process and a 30-item management skills assessment in the Leadership Practices Inventory (LPI) questionnaire[13, 14]. Managers completed the LPI self-assessment, and the clinic staff also shared their assessment of the managers’ abilities using the LPI tool. This produced a 360-degree assessment of the PHC managers. Staff interviews also assessed the impact of managers on the clinic environment and decision-making processes. PHC managers (n=20) and 100 staff were interviewed immediately after the training.

### Outcome Measures and Analysis Plan

The study prospectively assessed several process and outcome indicators (***Table 1***) during the 12-month in-training period. Process indicators included program completion rates and participant retention.

**Table 1.** Realised key performance indicators aligned with planned data sources and analysis plan.

| **Planned Assessment components** | **Measure** | **Instruments** | **Data source** |
| --- | --- | --- | --- |
| **Clinic Manager and staff** |  |  |  |
| Primary Outcome | Managerial Competence: Improvements in leadership and management skills of PHC managers, as evidenced by self-assessments and staff feedback. | Leadership practices inventory (LPI) 360 and Kotter's change model enquiry in study survey | Quantitative |
| Secondary Outcomes | Staff Performance: Staff morale and stress | Study survey | Quant/ qual |
|  | Staff experience of change introduction and management (Kotter's change model) | Study survey | Quantitative |

### Data analysis for Quantitative outcomes

#### Manager Competency: Leadership Practice Inventory (LPI)

The leadership practices of clinic managers were evaluated from the perspective of their staff using the 30-item LPI. Staff members rated their managers on a five-point Likert scale, ranging from 1 (rarely or never) to 5 (frequently or always), across 30 behavioural statements. These items were aggregated into five pre-determined distinct subscales, each comprising six items, to measure the core domains of the LPI tool: (1) Challenging the Process, (2) Inspiring a Shared Vision, (3) Enabling Others to Act, (4) Modelling the Way, and (5) Encouraging the Heart. For each domain, a continuous total score was generated by summing the relevant items, resulting in a potential score range of 6 to 30.

To facilitate qualitative interpretation and comparative analysis, these continuous scores were further transformed into categorical and binary variables. First, scores were grouped into three levels: “Low” (6–10), “Medium” (11–20), and “High” (21–30), representing the perceived frequency of leadership behaviours. Finally, these categories were collapsed into a binary indicator to distinguish between managers who demonstrated high-frequency leadership (scores of 21–30) and those perceived to demonstrate these behaviours with low-to-moderate frequency (scores of 6–20). This dual approach enabled both a granular assessment of leadership intensity and a robust binary classification for subsequent regression modelling.

#### Organisational readiness for change – Kotter’s model

We created a scale based on Kotter’s models to assess organisational readiness for change (***Table 2***). The scale has 10 items, each representing a specific stage of Kotter’s model. It uses a 5-point Likert scale (1 = Strongly Disagree to 5 = Strongly Agree). The clinic staff questionnaire gauged perceived readiness for service improvement initiatives, with perceptions anchored at the clinic level (clinic manager/clinic staff) rather than at the district leadership level, which was addressed in the manager questionnaire.

**Table 2.**
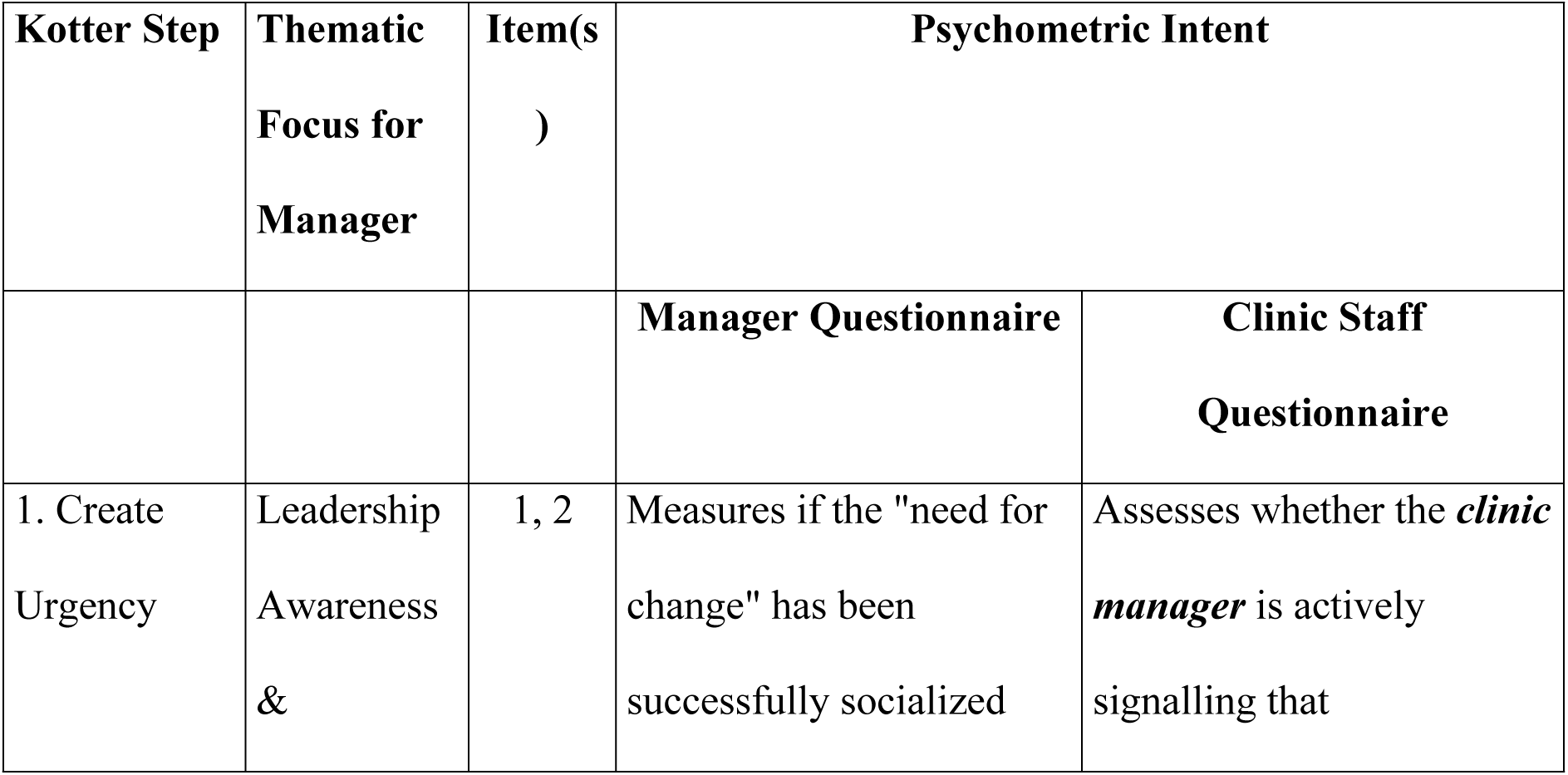
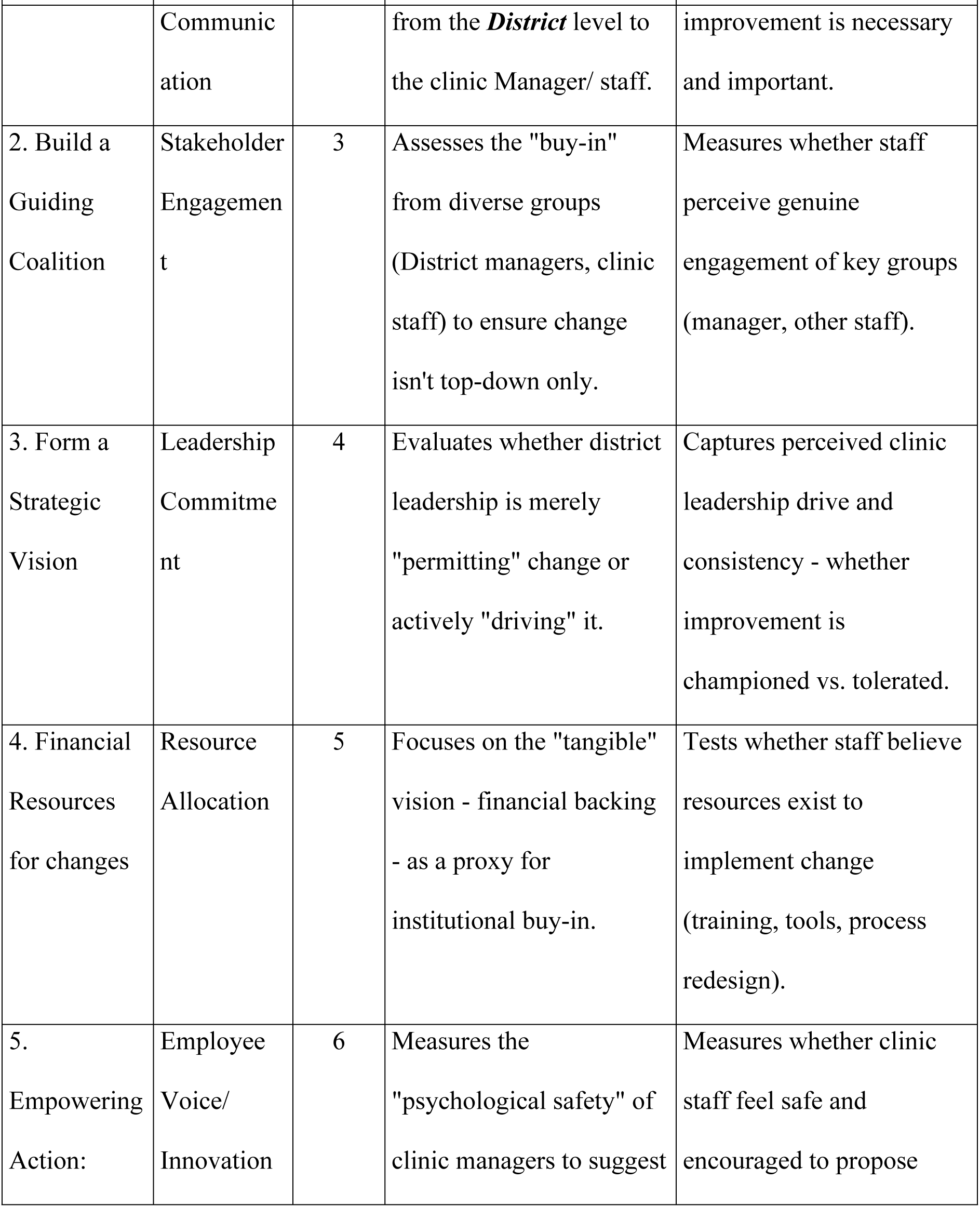
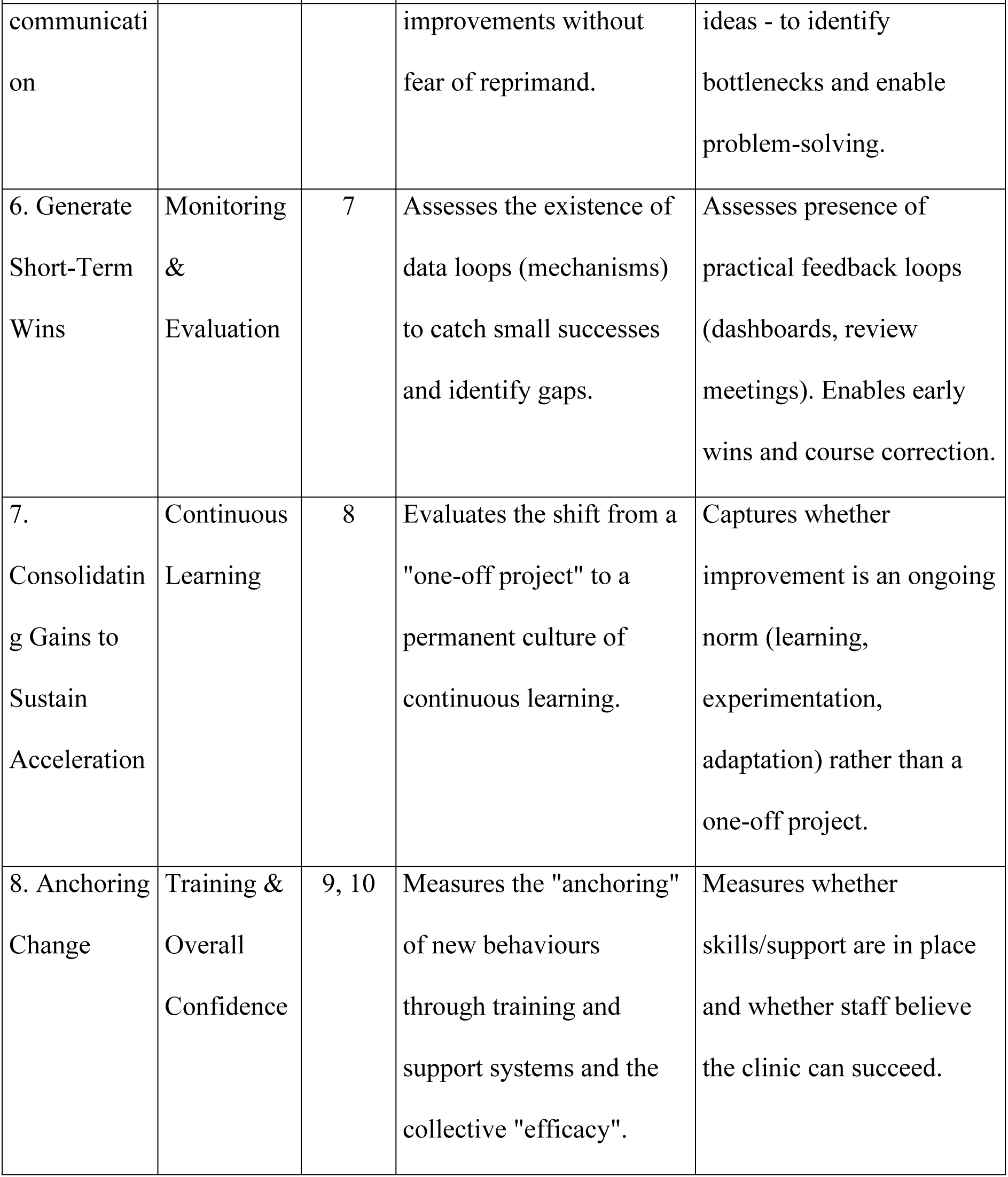
Integrated Change Management Readiness Framework — Mapping Kotter’s 8-Step Model across District/Manager and Clinic Staff Perspectives.

#### Clinic Manager subscales

Principal Component Factor (PCF) with an oblique (Direct Oblimin) rotation, was used to group related items into meaningful subscales, with factors retained based on eigenvalues (>=1). Following PCF analysis the survey items clustered into two distinct thematic subscales, collectively explaining 70.1% of the total variance. The first subscale, ***District Support and Structural Readiness***, comprised four items (district acknowledgement of the need for improvement, clear communication from the district, active engagement of stakeholders, and the sufficiency of allocated financial resources). This subscale captures the “external” enabling environment, focusing on whether the broader health system provides the necessary strategic material resources required for success (Cronbach’s alpha (α) = 0.78). The second subscale, ***Internal Clinic Leadership and Cultural Readiness***, comprised six items (clinic leadership commitment, encouragement of employee ideas, establishment of monitoring mechanisms, fostering a culture of continuous learning, provision of adequate training, and overall perceived preparedness). This subscale reflects the manager’s perception of the district’s support for change - specifically the extent to which the internal team is empowered, monitored, and culturally aligned with improvement goals (α = 0.90).

### Clinic Staff Subscales

In the PCF analysis, the items clustered into two main thematic subscales, explaining 65% of the variance. The first subscale, ***Relational Leadership and Strategic Engagement***, comprised six items (clinic manager acknowledgement of need for improvement, clear communication, active engagement of stakeholders, leadership commitment to driving improvement, encouragement for staff to share ideas, and establishment of mechanisms to monitor service quality). This subscale captures the extent to which staff perceive visible leadership, inclusive engagement, and an enabling environment for improvement (α = 0.83). The second subscale, ***Institutionalised Capability and Learning Culture***, comprised three items (culture of continuous learning and innovation, adequate training and support to implement changes, and overall perceived preparedness of the clinic to enhance services). This subscale reflects perceived organisational capability and the extent to which change is likely to be sustained through skills, support, and collective efficacy. Internal consistency for this three-item subscale was acceptable (α = 0.73).

#### Manager Stress

In addition to the Kotter-based organisational readiness measures, the survey included a standardised measure of manager occupational stress to assess whether MDP participation was associated with changes in perceived job demands and work conditions. The 13-item Job Stress Scale was designed to assess multiple dimensions of occupational stress among health facility managers, including workload demands, autonomy and control, skill utilisation, work environment, and overall job appraisal. Items were reverse-coded where appropriate so that higher scores consistently reflected lower stress or more favourable working conditions (e.g., lower workload burden, better environment, greater control). The overall job stress scale of 13 items demonstrated low reliability (α = 0.51), indicating that the full set of items did not function as a unidimensional construct in this sample. The PCF analysis retained two main subscales with reasonable internal consistency, explaining 39.91% of the variability.

The first dimension reflects ***Job Meaning and Environmental Quality***, capturing the manager’s overall appraisal of their role and the physical conditions of their workplace. This subscale comprises five items: the perceived technical difficulty and knowledge requirements of the position, the level of physical labour involved, the personal suitability of the job, the perceived worth of the work, and the quality of the physical environment (including noise, lighting, and ventilation) (α = 0.68).

The second dimension represents ***Autonomy and Control***, capturing the manager’s perceived decision latitude and flexibility in work organisation. This subscale comprises four items: the ability to have a say in what happens at work, control over one’s own work pace, the capacity to influence workplace policies and methods, and the cognitive demand of constantly thinking about work during the day. Within the framework of job stress theory, this cluster reflects the degree of professional agency and the ability to buffer workplace pressures through personal discretion (α = 0.69).

#### Quantitative Analysis Approach

The analysis used a generalised linear modelling (GLM) to estimate the intervention effect on dichotomous (binary) outcomes, accounting for the matched study design and clustering. Specifically, a GLM with a binomial family and identity link was fitted to directly estimate absolute risk differences with intervention status as the primary exposure. The model adjusted for manager experience as a continuous covariate and included matched-pair fixed effects. Standard errors were computed using cluster-robust variance estimation to account for within-clinic correlation. Following model estimation, the intervention coefficient was transformed to a percentage-point scale, yielding the adjusted risk difference expressed as a percent point (PP) difference in outcome between intervention and comparison study arms, with 95% confidence intervals and p-values.

## RESULTS

The baseline characteristics of clinic managers and staff across are presented in **Table 3**. Among clinic managers, the sex distribution was identical between the comparison and intervention arms, with 90.0% of managers in both groups being female. The median age of managers in the intervention arm was 44.5 years (IQR: 39-52), compared with 53.5 years (IQR: 51-56) in the comparison arm, and a statistically significant age imbalance. Additionally, there was a significant difference in professional tenure: 30.0% of comparison managers had served for 10 or more years in their current positions, whereas none of the intervention managers had equivalent tenure. In contrast to the managers, the clinic staff showed generally well-balanced characteristics across the study arms. The majority of staff in both groups were female, 72.0% in the comparison arm and 81.8% in the intervention arm. Median ages were similar at 43 years for comparison staff and 41 years for intervention staff, and professional experience was comparable across groups.

**Table 3.**
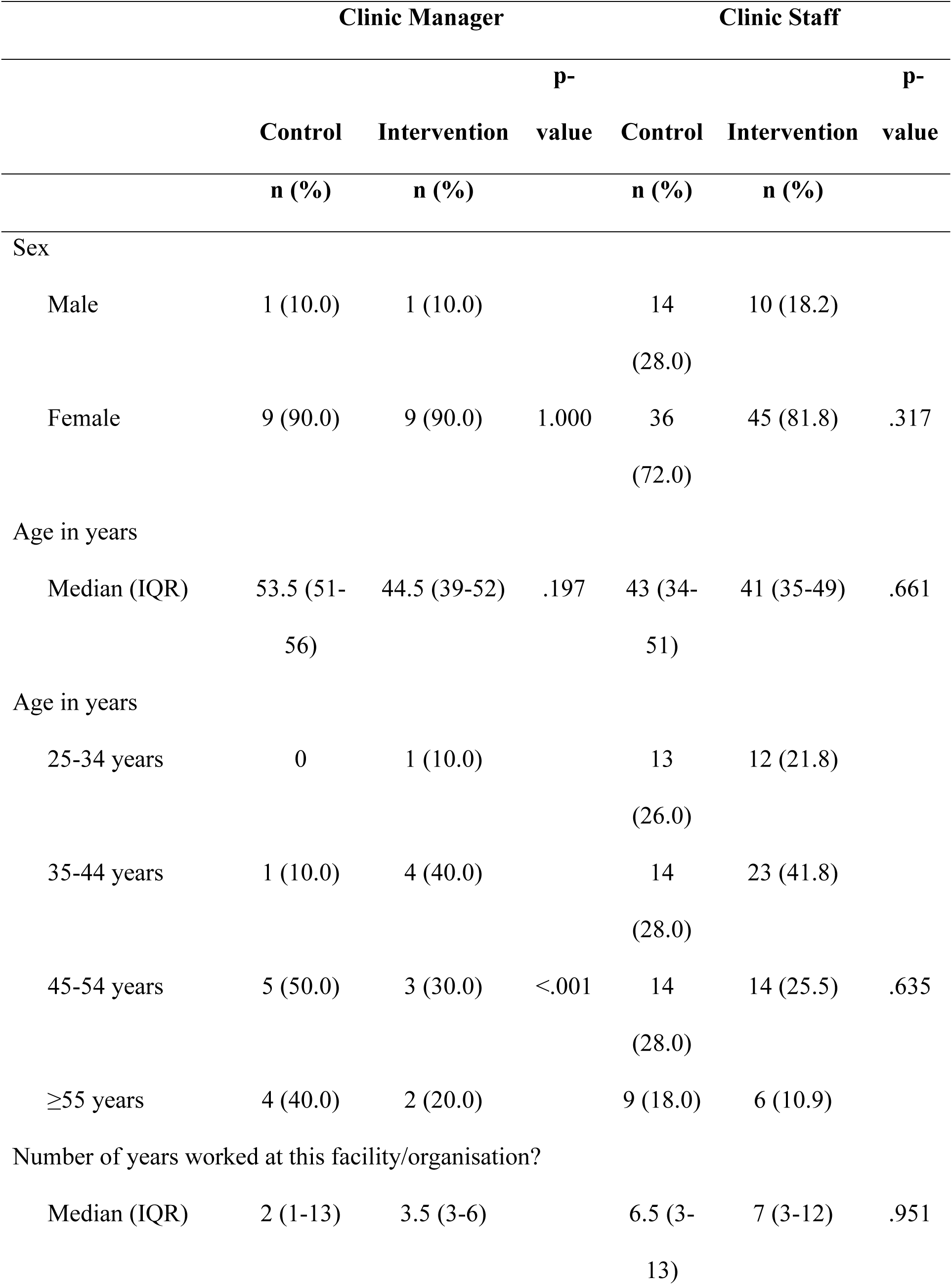
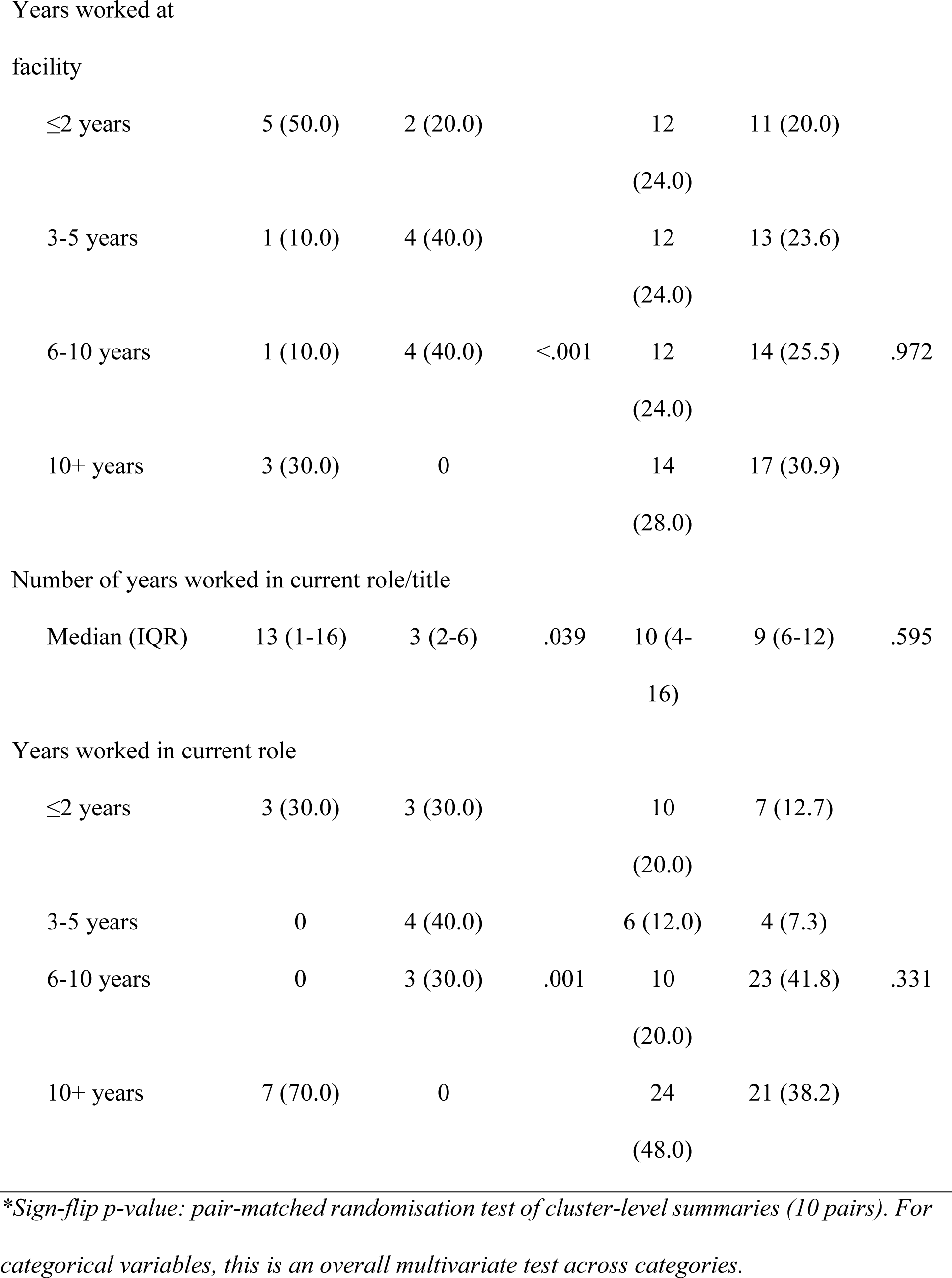
Characteristics of clinic managers and staff.

|  | Clinic Manager |  |  | Clinic Staff |  |  |
| --- | --- | --- | --- | --- | --- | --- |
|  | Control | Intervention | p-value | Control | Intervention | p-value |
|  | n (%) | n (%) |  | n (%) | n (%) |  |
| Sex |  |  |  |  |  |  |
| Male | 1 (10.0) | 1 (10.0) |  | 14 (28.0) | 10 (18.2) |  |
| Female | 9 (90.0) | 9 (90.0) | 1.000 | 36 (72.0) | 45 (81.8) | .317 |
| Age in years |  |  |  |  |  |  |
| Median (IQR) | 53.5 (51-56) | 44.5 (39-52) | .197 | 43 (34-51) | 41 (35-49) | .661 |
| Age in years |  |  |  |  |  |  |
| 25-34 years | 0 | 1 (10.0) |  | 13 (26.0) | 12 (21.8) |  |
| 35-44 years | 1 (10.0) | 4 (40.0) |  | 14 (28.0) | 23 (41.8) |  |
| 45-54 years | 5 (50.0) | 3 (30.0) | <.001 | 14 (28.0) | 14 (25.5) | .635 |
| ≥55 years | 4 (40.0) | 2 (20.0) |  | 9 (18.0) | 6 (10.9) |  |
| Number of years worked at this facility/organisation? |  |  |  |  |  |  |
| Median (IQR) | 2 (1-13) | 3.5 (3-6) |  | 6.5 (3-13) | 7 (3-12) | .951 |
| ≤2 years | 5 (50.0) | 2 (20.0) |  | 12<br>(24.0) | 11 (20.0) |  |
| 3-5 years | 1 (10.0) | 4 (40.0) |  | 12<br>(24.0) | 13 (23.6) |  |
| 6-10 years | 1 (10.0) | 4 (40.0) | <.001 | 12<br>(24.0) | 14 (25.5) | .972 |
| 10+ years | 3 (30.0) | 0 |  | 14<br>(28.0) | 17 (30.9) |  |
| Number of years worked in current role/title |  |  |  |  |  |  |
| Median (IQR) | 13 (1-16) | 3 (2-6) | .039 | 10 (4-16) | 9 (6-12) | .595 |
| Years worked in current role |  |  |  |  |  |  |
| ≤2 years | 3 (30.0) | 3 (30.0) |  | 10<br>(20.0) | 7 (12.7) |  |
| 3-5 years | 0 | 4 (40.0) |  | 6 (12.0) | 4 (7.3) |  |
| 6-10 years | 0 | 3 (30.0) | .001 | 10<br>(20.0) | 23 (41.8) | .331 |
| 10+ years | 7 (70.0) | 0 |  | 24<br>(48.0) | 21 (38.2) |  |

*\*Sign-flip p-value: pair-matched randomisation test of cluster-level summaries (10 pairs). For categorical variables, this is an overall multivariate test across categories.*

The results of the clinic staff assessment of their managers and work environment, as presented in **Table**, indicate that while the five core domains of the LPI remained largely unchanged, there were meaningful improvements in specific managerial attributes. For the standard LPI domains—Challenging the Process, Inspiring a Shared Vision, Enabling Others to Act, Modelling the Way, and Encouraging the Heart —the adjusted % risk differences were small and not statistically significant. In these areas, staff in both the comparison and intervention arms reported high levels of leadership practices, ranging from 68.0% to 81.8%, suggesting a high perceived leadership that was not further shifted by the intervention.

In contrast, the intervention was associated with significant positive shifts in staff perceptions of their manager’s relational skills and supporting staff readiness to organisational changes. The proportion of staff reporting high scores for their manager’s relational and communication practices was 72.7% in the intervention arm compared to 64.0% in the comparison arm, resulting in a significant adjusted risk difference of 22.0% (95% CI: 6.1, 37.8; p=.007). Similarly, although the raw percentage of high scores for manager training and readiness (institutionalised capability and learning culture) was slightly lower in the intervention group (69.1% vs 72.0%), the adjusted model, which accounted for baseline imbalances in manager age and tenure, revealed a significant positive intervention effect. The adjusted risk difference for training and readiness was 21.3% (95% CI: 0.6, 42.0; p=0.043). These findings suggest that after adjusting for the fact that intervention managers were generally younger and less experienced, the intervention significantly improved staff perceptions of their managers’ communication effectiveness and their overall readiness for the role.

**Table 4.**
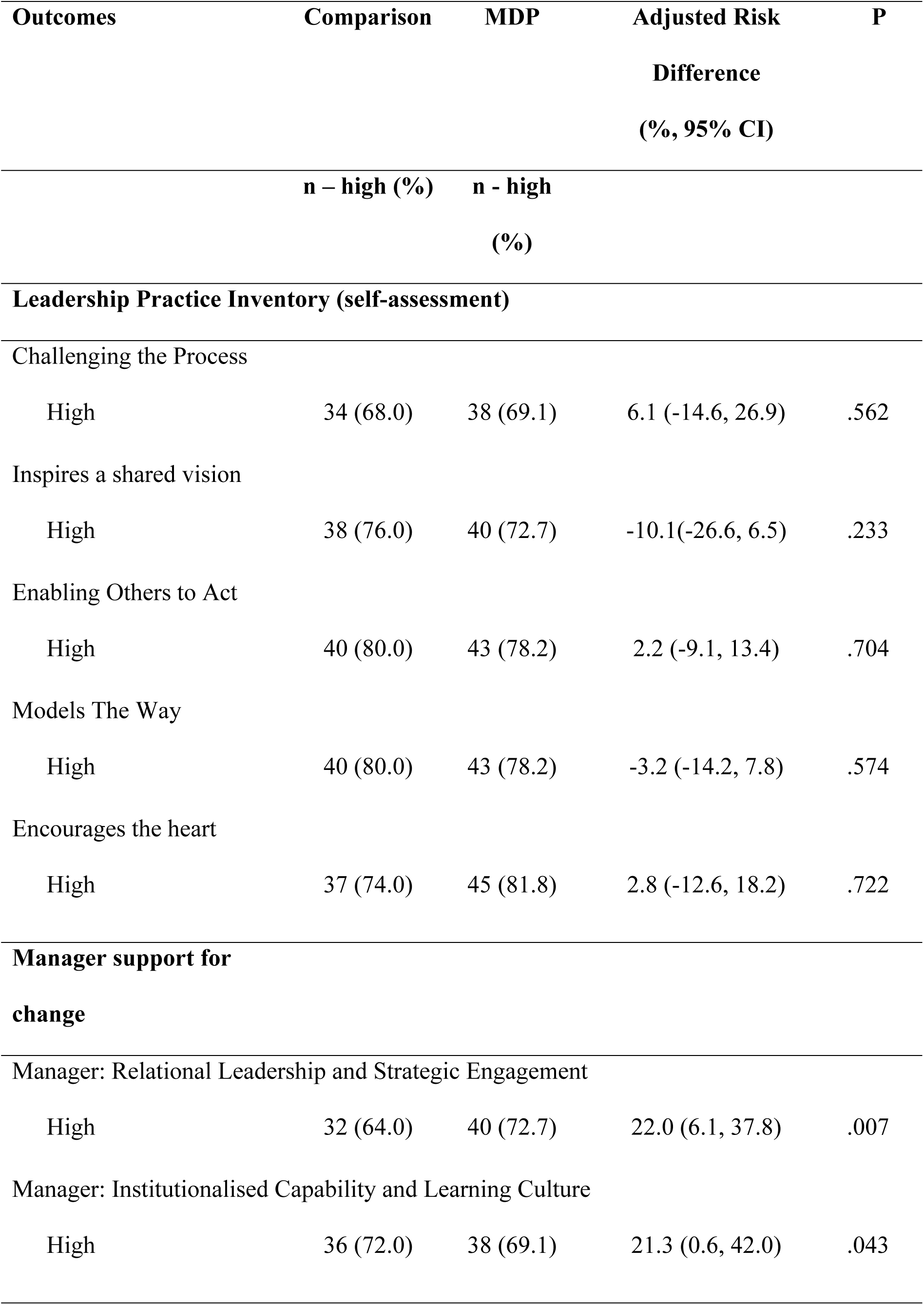
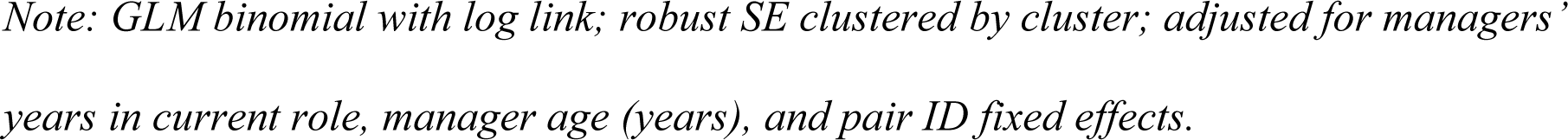
Leadership Practices Inventory - clinic staff assessment of manager and environment.

| Outcomes | Comparison | MDP | Adjusted Risk<br>Difference<br>(%, 95% CI) | P |
| --- | --- | --- | --- | --- |
|  | n – high (%) | n - high<br>(%) |  |  |
| **Leadership Practice Inventory (self-assessment)** |  |  |  |  |
| Challenging the Process |  |  |  |  |
| High | 34 (68.0) | 38 (69.1) | 6.1 (-14.6, 26.9) | .562 |
| Inspires a shared vision |  |  |  |  |
| High | 38 (76.0) | 40 (72.7) | -10.1(-26.6, 6.5) | .233 |
| Enabling Others to Act |  |  |  |  |
| High | 40 (80.0) | 43 (78.2) | 2.2 (-9.1, 13.4) | .704 |
| Models The Way |  |  |  |  |
| High | 40 (80.0) | 43 (78.2) | -3.2 (-14.2, 7.8) | .574 |
| Encourages the heart |  |  |  |  |
| High | 37 (74.0) | 45 (81.8) | 2.8 (-12.6, 18.2) | .722 |
| **Manager support for**<br>**change** |  |  |  |  |
| Manager: Relational Leadership and Strategic Engagement |  |  |  |  |
| High | 32 (64.0) | 40 (72.7) | 22.0 (6.1, 37.8) | .007 |
| Manager: Institutionalised Capability and Learning Culture |  |  |  |  |
| High | 36 (72.0) | 38 (69.1) | 21.3 (0.6, 42.0) | .043 |

*Note: GLM binomial with log link; robust SE clustered by cluster; adjusted for managers’* *years in current role, manager age (years), and pair ID fixed effects.*

**Table** summarises managers’ self-assessed leadership practices, perceived district support, and job stress by study arm, with models adjusted for years in the current role. Across the LPI self-assessment domains, managers reported uniformly high scores in both arms, including a ceiling effect for “Challenging the Process” where 100% of managers in both groups were classified as high. For the remaining leadership domains, the intervention arm generally had a higher proportion of high scores than the comparison arm, but the adjusted risk differences were not statistically significant; estimated effects ranged from approximately 19.8% to 21.7% with wide confidence intervals spanning zero (all p-values >.100).

In contrast, managers in the intervention arm reported significantly higher levels of perceived district support for change. The proportion reporting high internal leadership and cultural readiness was 70.9% in the intervention group versus 62.0% in the comparison group, corresponding to an adjusted risk difference of 22.1% (95% CI 14.0 to 30.3; p<.001). Similarly, high ratings for district leadership and resource availability were more common in the intervention arm (47.3% vs 36.0% for the comparison clinics), with an adjusted risk difference of 28.1% (95% CI 15.6 to 40.6; p<.001).

**Table 5.**
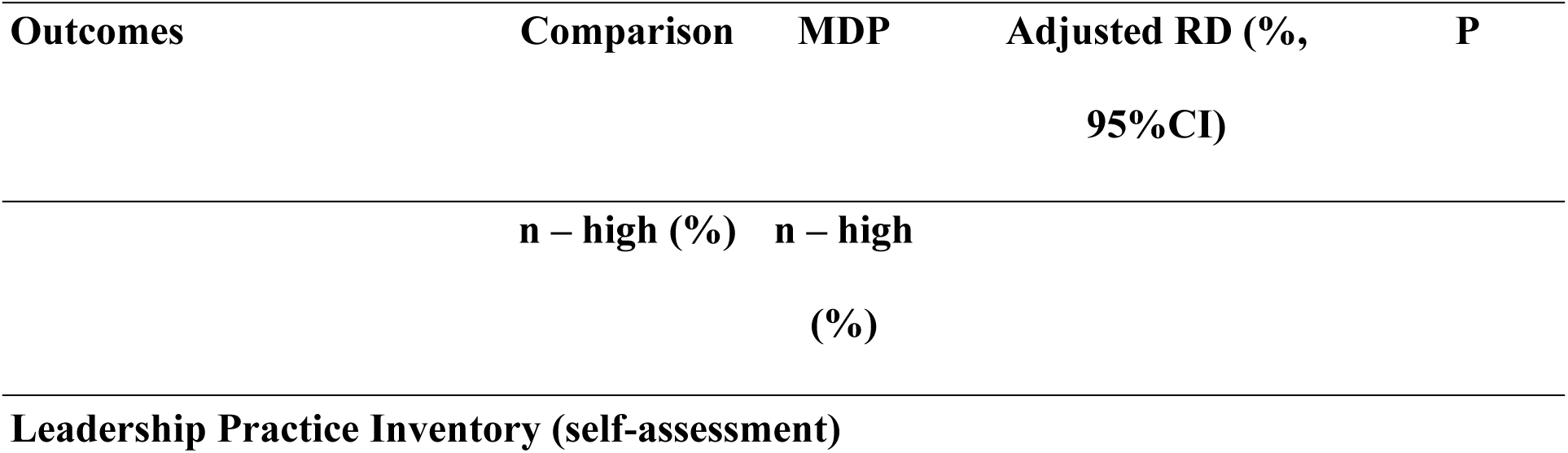
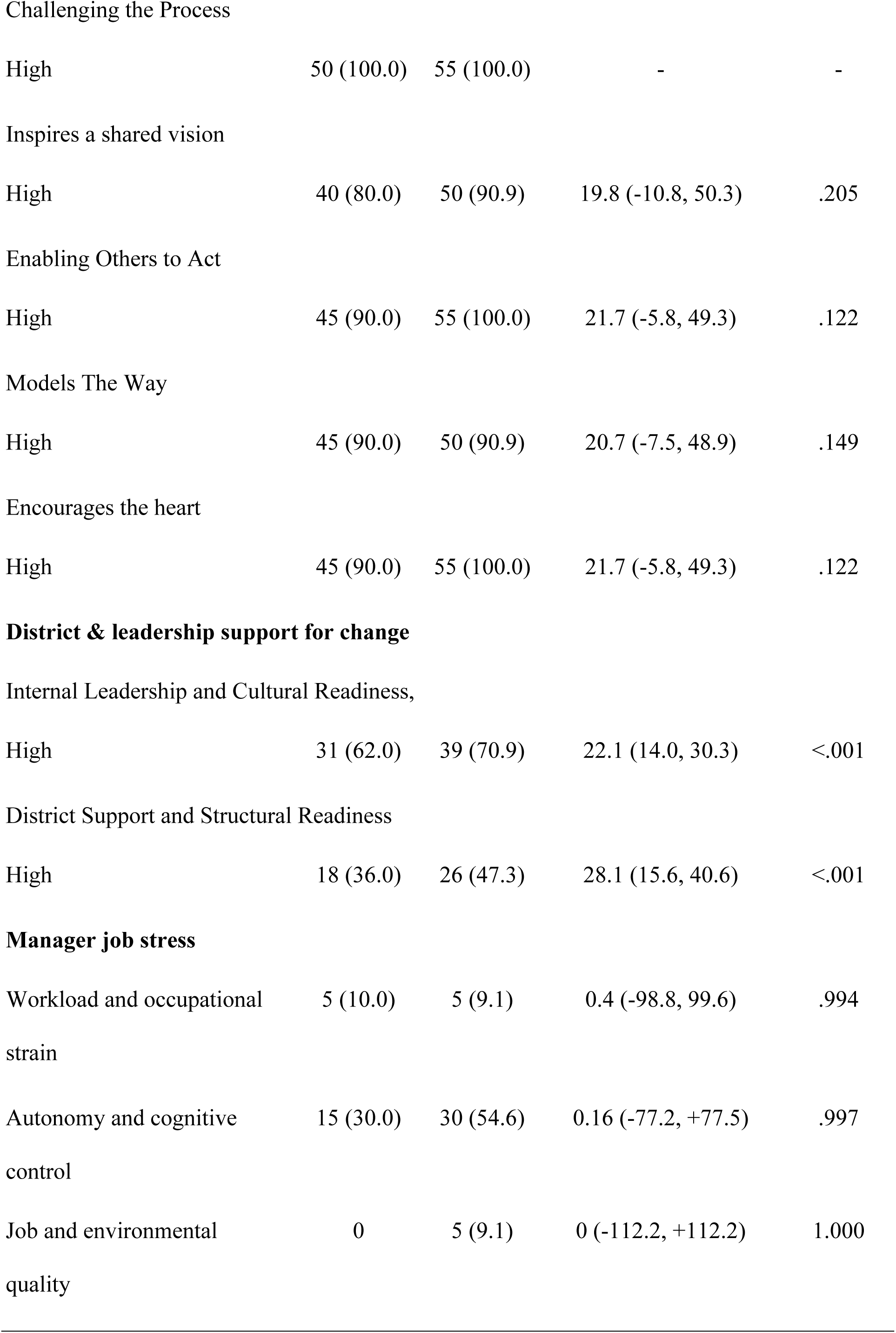
Manager competency self-assessment.

| Outcomes | Comparison | MDP | Adjusted RD (%,<br>95%CI) | P |
| --- | --- | --- | --- | --- |
|  | n – high (%) | n – high<br>(%) |  |  |
| Leadership Practice Inventory (self-assessment) |  |  |  |  |
| Challenging the Process |  |  |  |  |
| High | 50 (100.0) | 55 (100.0) | - | - |
| Inspires a shared vision |  |  |  |  |
| High | 40 (80.0) | 50 (90.9) | 19.8 (-10.8, 50.3) | .205 |
| Enabling Others to Act |  |  |  |  |
| High | 45 (90.0) | 55 (100.0) | 21.7 (-5.8, 49.3) | .122 |
| Models The Way |  |  |  |  |
| High | 45 (90.0) | 50 (90.9) | 20.7 (-7.5, 48.9) | .149 |
| Encourages the heart |  |  |  |  |
| High | 45 (90.0) | 55 (100.0) | 21.7 (-5.8, 49.3) | .122 |
| Internal Leadership and Cultural Readiness, |  |  |  |  |
| High | 31 (62.0) | 39 (70.9) | 22.1 (14.0, 30.3) | <.001 |
| District Support and Structural Readiness |  |  |  |  |
| High | 18 (36.0) | 26 (47.3) | 28.1 (15.6, 40.6) | <.001 |
| **Manager job stress** |  |  |  |  |
| Workload and occupational strain | 5 (10.0) | 5 (9.1) | 0.4 (-98.8, 99.6) | .994 |
| Autonomy and cognitive control | 15 (30.0) | 30 (54.6) | 0.16 (-77.2, +77.5) | .997 |
| Job and environmental quality | 0 | 5 (9.1) | 0 (-112.2, +112.2) | 1.000 |

### Adjusted for the number of years in the current role

For manager job stress outcomes, there was no evidence of differences between arms. High/difficult workload was rare and essentially identical between groups (10.0% vs 9.1%), and the adjusted risk difference was near zero with an extremely wide confidence interval. The “control of time/pace” and “job quality/fit” stress measures also did not show statistically significant differences, with estimates close to zero and very imprecise confidence intervals, reflecting limited outcome variation

## DISCUSSION

This evaluation of the MDP provides imperative evidence about the program’s impact on primary health care. Overall, the program showed limited short-term effects on leadership competencies, as we found no significant differences between managers trained under the MDP and those who were not, and this was based on the staff’s perspectives. Both reported high levels of leadership practices, but this was not further improved by the intervention. The findings align with previous findings that observable leadership takes longer to reflect than organisational culture, and that longitudinal follow-up studies are a more appropriate way to measure leadership development [19]. This is motivated by the idea that leadership development is an ongoing journey and requires prolonged practice and mentorship [20].

However, the staff in the intervention group reported notable improvements in organisational change readiness within PHC services, specifically the relational skills and supporting staff readiness for the organisational change. These findings are consistent with previous studies that have shown that leadership is important in creating a climate of psychological safety that enables team members to engage in learning behaviours and promotes teamwork [21–23]. These are key elements of Kotter’s early change steps. Additionally, the quality of manager-staff relationships has been shown to directly shape staff motivation, job satisfaction, and willingness to engage with improvements of services within facilities [24, 25].

Regarding the LPI self-assessment domains, managers reported uniformly high scores in both arms. It has been shown that while managers often rate their own communication competency highly, their supervisors and subordinates, however, consistently rate them lower, pointing to a persistent gap between how managers perceive themselves and how they are experienced by the people they lead [26]. The present intervention appears to have the potential to meaningfully close that gap.

Additionally, managers in the intervention arm were significantly more likely to report feeling supported by the district, with high scores for leadership and cultural readiness. Even more notably, high ratings for district leadership and resource availability were substantially more common in the intervention group. Studies have shown that facility managers and staff can only meaningfully engage with and sustain organisational change when they feel that the district is visibly behind them, providing leadership, resources, and a culture that enables rather than obstructs implementation [27]. The fact that a facility-level intervention produced improvements in perceived district support suggests it changed not only how managers led within their clinics, but how they engaged with the system around them.

Regarding the job stress, there were no differences found between managers in the intervention arm compared to the control arm. This is in contrast to previous studies that reported increased awareness and accountability following the training, whereby a manager becomes more aware and recognises the weaknesses and their accountability burdens[28]. PHC managers frequently experience high stress levels, also due to having to constantly juggle clinical duties, administrative oversight, and human resource challenges, often with insufficient support. Managers often take on clinical duties at the expense of their managerial roles, contributing to frustration and stress in their work environment[24]. In addition, heavy workloads and insufficient time to fulfil administrative tasks have also been shown to contribute to increased stress, anxiety, and depression among facility managers [29].

Strong leadership is the foundation for the effective implementation of South Africa’s NHI, which will require high performing PHC facilities capable of coordinating integrated, people-centred care [30, 31]. Although the MDP’s immediate effects on leadership competencies appear limited, improvements in organisational culture and change readiness are promising, as these are essential precursors for sustained transformation. The MDP may therefore have a greater impact over time as trained managers accumulate experience and engage in ongoing mentorship for longer periods of time.

Our evaluation had strengths and some limitations. The strengths of this evaluation include its matched cluster design, multisource 360-degree leadership assessment, and integration of both quantitative and qualitative inputs. However, the study is limited by its small sample size, potential selection bias which is associated with purposive enrolment of intervention facilities, and the short lag time between training and evaluation for some of the facility managers. Leadership behaviour change is inherently longitudinal and may require sustained follow-up beyond the timeframe of most trials to become reliably detectable [32]. Future follow-up assessments with larger cohorts will be essential.

## CONCLUSIONS

This early evaluation shows that while the MDP did not significantly improve observable leadership behaviours in the short term, it positively influenced organisational readiness for change and staff perceptions of leadership. These findings underscore the value of structured leadership training but also highlight the need for longer-term follow-up, ongoing coaching, and alignment with broader system reforms. As South Africa progresses toward NHI implementation, strengthening PHC leadership through programmes like the MDP will be critical in enabling effective and impartial service delivery.

## Data Availability

The de-identified data supporting the findings of this study are available from the corresponding author.

## ACKNOWLEDGEMENTS

We extend our gratitude to the data collection team (Zanele Walaza, Zolani Mtwa, Avile Fatyela) for their diligent support during the study implementation. Additionally, our sincere thanks go to the City of Cape Town and Western Cape Department of Health and the staff of participating clinics for accommodating the study. We also thank patients attending these clinics for their willingness to participate and share their valuable

